# Pre-vaccination transcriptomic profiles of immune responders to the MUC1 peptide vaccine for colon cancer prevention

**DOI:** 10.1101/2024.05.09.24305336

**Authors:** Cheryl M. Cameron, Vineet Raghu, Brian Richardson, Leah L. Zagore, Banumathi Tamilselvan, Jackelyn Golden, Michael Cartwright, Robert E. Schoen, Olivera J. Finn, Panayiotis V. Benos, Mark J. Cameron

**Affiliations:** Department of Nutrition, Case Western Reserve University, Cleveland, OH; Department of Computer Science, University of Pittsburgh, Pittsburgh, PA; Massachusetts General Hospital, Harvard Medical School, Cambridge, MA; Department of Population and Quantitative Health Sciences, Case Western Reserve University, Cleveland, OH; Division of Gastroenterology, Hepatology and Nutrition, University of Pittsburgh, Pittsburgh, PA; Department of Immunology, University of Pittsburgh, Pittsburgh, PA; Department of Epidemiology, University of Florida, Gainesville, FL; Department of Computational and Systems Biology, University of Pittsburgh, Pittsburgh, PA

**Author notes:** Contributed equally (Cheryl M. Cameron, Vineet Raghu, Brian Richardson).

**Keywords:** colon cancer, colorectal adenoma, cancer vaccine, transcriptomics, MUC1, serological response

## Abstract

Self-antigens abnormally expressed on tumors, such as MUC1, have been targeted by therapeutic cancer vaccines. We recently assessed in two clinical trials in a preventative setting whether immunity induced with a MUC1 peptide vaccine could reduce high colon cancer risk in individuals with a history of premalignant colon adenomas. In both trials, there were immune responders and non-responders to the vaccine. Here we used PBMC pre-vaccination and 2 weeks after the first vaccine of responders and non-responders selected from both trials to identify early biomarkers of immune response involved in long-term memory generation and prevention of adenoma recurrence. We performed flow cytometry, phosflow, and differential gene expression analyses on PBMCs collected from MUC1 vaccine responders and non-responders pre-vaccination and two weeks after the first of three vaccine doses. MUC1 vaccine responders had higher frequencies of CD4 cells pre-vaccination, increased expression of CD40L on CD8 and CD4 T-cells, and a greater increase in ICOS expression on CD8 T-cells. Differential gene expression analysis revealed that iCOSL, PI3K AKT MTOR, and B-cell signaling pathways are activated early in response to the MUC1 vaccine. We identified six specific transcripts involved in elevated antigen presentation, B-cell activation, and NF-κB1 activation that were directly linked to finding antibody response at week 12. Finally, a model using these transcripts was able to predict non-responders with accuracy. These findings suggest that individuals who can be predicted to respond to the MUC1 vaccine, and potentially other vaccines, have greater readiness in all immune compartments to present and respond to antigens. Predictive biomarkers of MUC1 vaccine response may lead to more effective vaccines tailored to individuals with high risk for cancer but with varying immune fitness.

## Introduction

Self-antigens abnormally expressed in tumors, known as non-viral cancer-associated antigens, have been extensively tested over the last three decades as antigens in therapeutic cancer vaccines (1-3). In preclinical studies, an immune response to these antigens can prevent cancer growth without causing toxicity. In humans, preexisting immunity to some such antigens correlates with better disease outcome or reduced risk of cancer recurrence (4). Nevertheless, therapeutic vaccines utilizing these antigens have had low immunogenicity and no clinical efficacy. This has been attributed to the presence of many immunosuppressive influences in the tumor microenvironment (5, 6). MUC1 is a cancer-associated antigen that has been effective as a vaccine in preclinical animal models but showed limited immunogenicity and efficacy as a therapeutic vaccine in clinical trials in colon, breast, pancreas, prostate and lung cancer (7-11). Hypothesizing that the major difference between the outcome of the vaccine in preclinical models and clinical trials is the high level of immune suppression in cancer patients, we began to develop models and MUC1 vaccines for cancer prevention in patients at risk; before immune suppression develops. As MUC1 is expressed on early premalignant lesions as well as cancer, we chose to study immunogenicity, safety and potential efficacy of this vaccine in the preventative setting in individuals with a history of colonic polyps that increases their risk of colon cancer (12).

From 2008 to 2012, we conducted a single arm trial (NCT-007773097) (13) in 41 individuals. Forty-three percent (43%) of vaccinated participants responded to the vaccine as measured by production of anti-MUC1 IgG at week 12 post vaccination (vaccine responders), and 57% did not respond (vaccine non-responders). From 2015-2020, we conducted the second study, a randomized, double-blind placebo-controlled multi-center efficacy trial of the same MUC1 vaccine in the setting of newly diagnosed advanced adenomas in 110 individuals (NCT-02134925) (14). Twenty-seven percent (27%) of the vaccinated participants responded to the vaccine. In addition to the immune response, in this trial we evaluated adenoma recurrence by follow-up colonoscopy ≥1 year from the start of vaccination. In vaccine responders, adenoma recurrence was reduced by 38% compared to non-responders and placebo controls. Predictable factors such as gender, age, and HLA-type were not significantly different between vaccine responders and non-responders. It became important to understand why some individuals mounted a potentially protective immune response, while others did not, having the same diagnosis.

In this study, we analyzed PBMC samples collected from both trials at baseline (pre-vaccination) and 2 weeks post-first of 3 vaccines (week 0, week 2 and week 10) from vaccine responders and non-responders and identified comprehensive gene and pathway biomarkers related to vaccine response. We discovered that several key T-and B-cell cellular proliferation and stress pathways were enriched in responders, while oxidative phosphorylation and DNA damage response and repair pathways were enriched in non-responders. Responders had higher frequencies of CD4 cells at baseline, with higher activation and/or costimulatory signaling in CD8 and CD4 T-cells from baseline to week 2 in CD8 T-cells. Phosflow analysis revealed enhanced phosphorylation of B-cell signaling molecules and T-cell help targets in responders at baseline and a significant increase in NFκB phosphorylation in B-cells at week 2. Lastly, we applied graphical modeling approaches (15-17) to this data and built a regression model to discriminate future responders and non-responders via their predicted and actual IgG response at week 12.

## Materials and Methods

### PBMC collection

PBMC samples from patients with a history of, or with newly diagnosed, advanced colonic adenoma and at high risk for colon cancer were collected as part of two clinical trials of a MUC1 vaccine registered at clinicaltrials.gov (NCT-007773097, NCT-02134925) (13, 14). The ethics committee/IRB of the following institutions gave ethical approval for this work: Mayo Clinic, Rochester MN; Kansas City Veterans Affairs Medical Center, Kansas City, KS; University of Pittsburgh Medical Center, Pittsburgh PA; University of Puerto Rico, San Juan PR; Thomas Jefferson University Hospital, Philadelphia PA; and Massachusetts General Hospital, Boston MA. All participants provided written informed consent. Blood samples were processed within 24 hours by the same individual, using the same protocol. Heparinized blood was layered on lymphocyte separation medium (MPbio) and centrifuged at 800 g for 10 min with lowest acceleration and deceleration speed. PBMC were collected from the interphase, washed twice, resuspended in 80% human serum and 20% DMSO, and stored in liquid nitrogen.

### RNA-Seq

PBMC samples were thawed, pelleted, and lysed in 350 uL of RLT with beta-mercaptoethanol. RNA was isolated using the RNeasy Mini kit (Qiagen). RNA quality was assessed with the Fragment Analyzer (Agilent) and its Standard Sensitivity RNA kit. Total RNA was normalized to 100 ng prior to random hexamer priming and libraries generated by the TruSeq Stranded Total RNA – Globin kit (Illumina). The resulting libraries were assessed on the Fragment Analyzer (Agilent) with the High Sense Large Fragment kit and quantified using a Qubit 3.0 fluorometer (Life Technologies. Medium depth sequencing (>30 million reads per sample) was performed with a HiSeq 2500 (Illumina) on a High Output, 125 base pair, Paired End run.

### Bioinformatic analysis

Raw demultiplexed fastq paired end read files were trimmed of adapters and filtered using the program skewer (18), discarding those with an average phred quality score <30 or a length <36. Trimmed reads were aligned to human reference genome GRCh38 using HISAT2(19) and sorted using SAMtools (20). Aligned reads were counted and assigned to gene meta-features using the program featureCounts (21) as part of the Subread package. Quality control, normalization and analysis were performed in R, using an in-house pipeline utilizing the limma-trend method for differential gene expression testing and the GSVA (22) library for gene set sample enrichment. Final differential gene expression lists were filtered to remove non-coding RNAs as well as LOC features. The datasets for this study can be found in the Gene Expression Omnibus (GEO) public database with the accession number pending.

### Flow cytometric analysis

For immune cell phenotyping and assessment of intracellular levels of bcl2, and phosphorylation of STAT3, erk1/2, NF-kB and MTORC targets, cells were first stained with Live/Dead Aqua (Invitrogen) followed by cocktails of monoclonal antibodies recognizing the following cell surface markers: CD4, CD8, CD45RA, CD27, CCR7, CD152, CD86, CD275, CD11c, CD56, CD16, CD19, CD3, HLA-DR, CD14, CD40, and CD11b. Cells were washed, fixed and permeabilized, then stained with antibodies specific for the following intracellular proteins: NFkB p65, erk1/2 (pT202/pY204), STAT3 (pY705), Akt1, pS6 (S235/236 & S240) and p4E-BP1 (T36/46). Cells were washed and fixed and events were collected on a BD ARIA-SORP instrument. A 15-minute incubation at 37C with recombinant human IL-6 (100ng/mL) (BD Pharmingen) was performed to induce NFκB signaling. After washing, cells were resuspended in staining buffer and sorted on an ARIA-SORP. Data was analyzed using FlowJo software (TreeStar).

### Predictive model for post-vaccination immune response

A detailed explanation of computational model development and evaluation can be found in the supplemental methods. Briefly, a LASSO logistic regression (23) was used to develop a prediction model for a binary outcome of response defined by the clinical trial endpoint (≥2-fold increase in IgG from baseline to week 12), using transcriptomic data measured two-weeks post-vaccination (Week 2 data). A Mixed Graphical Models (MGM) algorithm was used to infer a unidirectional graphical model followed by FCI-MAX to determine direction. All statistical analysis was performed in R.

### Statistics

Unless otherwise indicated, the Student’s t-test was used, with p ≤ 0.05 chosen as the level of significance.

## Results

### Vaccine responders and non-responders show differential gene expression in PBMCs pre-vaccination

Next-generation RNA-seq analysis was performed on PBMCs from 46 participants of the two trials, conducted in the same setting of advanced adenoma and with the identical vaccine and vaccination protocol. The vaccine, composed of 100μg of MUC1 peptide plus the polyICLC adjuvant Hiltonol®, was administered at week 0, 2, 10 and 52. We assayed PBMC samples collected at the time of the first injection (baseline) and 2 weeks later, at the time of the second injection in order to be able to define preexisting (at baseline) and early post-vaccination (week 2) signatures of response to MUC1 vaccination. Vaccine responders (R, n=13) in both trials were defined as having anti-MUC1 IgG levels at week 12 (after all three injections) at least two-fold higher than baseline. For some of our analyses we also classified responders by antibody levels into high responders (HR, anti-MUC1 IgG OD450 at 1:80 plasma dilution ≥0.4), low responders (LR, OD450 at 1:80 plasma dilution <0.4), and non-responders (NR) (no difference from baseline).

RNA-seq performed on PBMCs collected immediately pre-vaccination (baseline) revealed a total of 2,321 genes that were differentially expressed between all responders and all non-responders at baseline (Fig 1A). Within these genes, 1,337 showed increased transcript levels and 984 showed decreased levels. Among the top 50 differentially expressed genes by p-value (Fig 1B), 47 genes were upregulated in responders compared to non-responders. The upregulated genes were involved in transcriptional and epigenetic regulation, including multiple subunits of the SWI/SNF chromatin remodeling complex (ARID1A, ARID1B, and SMARCC2), as well as NCOA6, a multifunctional transcriptional coactivator and component of the Set1-like H3K4-methyltransferase complex ASCOM, all of which have been shown to play a role in the pathogenesis of cancer (24). Additional cancer-relevant transcriptional regulators higher in responders before vaccination include SP2, NFYC, AKNA, MYPOP, and ZNF652. CUX1 is a subunit of the NF-muNR repressor that binds to the matrix attachment regions of the immunoglobulin heavy chain enhancer and the TCR enhancer. The epigenetic regulator HCFC1 tethers Set and Sin3 histone modifying complexes together and was also higher in responders at baseline (25). We observed an increase in TRAPPC9, an activator of NFκB, and FAM168A, which is involved in the PI3K/AKT/NFκB signaling pathway. RRAGA was one of 3 downregulated genes among the top differentially expressed genes in responders and plays a role in regulating the mTORC1 complex (26). The presence of transcriptional and epigenetic regulators within the list of upregulated genes could explain the large number of significant changes in steady-state RNA levels we observed and suggests a differing global transcriptional program between responders and non-responders before vaccination. As these upstream regulators have the potential to broadly remodel the transcriptome, they may represent potent therapeutic targets.

**Figure 1:**
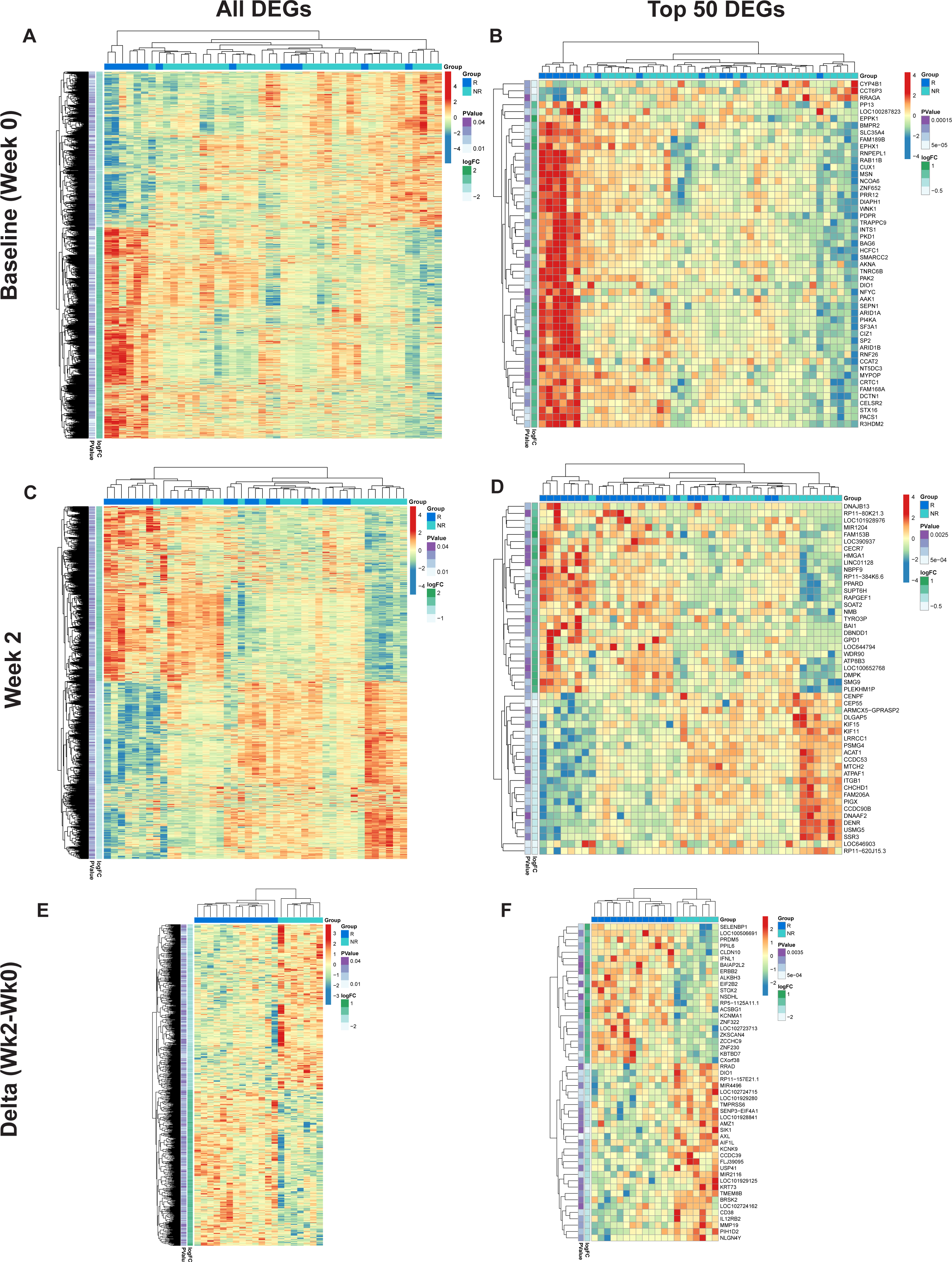
Differential gene expression pre- and post-MUC1 vaccination in responders and non-responders. Differentially expressed genes (DEGs) are shown for each time point and contrast in the left column, top 50 DEGs are shown in the right column. Group status is indicated in the row above the heatmap with responders (R) in dark blue, and non-responders (NR) in light blue. Z-scored normalized gene expression for each gene is displayed horizontally across all samples (diverging color scale legend on the upper right of each heatmap). Log2 fold-change and p values are indicated in the purple and green vertical columns, respectively. Unsupervised clustering of the samples is indicated by the black dendrogram at the top of heatmap, while clustering of the genes is indicated at the far left. Heatmaps showing all DEGs pre-vaccination (Baseline/Week 0) (A), top 50 DEGs pre-vaccination (Baseline/Week 0) (B), DEGs at Week 2 post-vaccination (C), top 50 DEGs at Week 2 post-vaccination (D), all genes demonstrating longitudinal changes at Week 2 vs. Week 0 (Delta Wk2-Wk0) (E), top 50 genes demonstrating longitudinal changes at Week 2 vs. Week 0 (Delta Wk2-Wk0) (F) in PBMCs from responders versus non-responders.

### Vaccine responders and non-responders show differences in gene expression in PBMCs at week 2 post-vaccination

At two weeks post-first injection, we found 1,887 genes differentially expressed in responders vs. non-responders, 934 genes upregulated and 953 genes downregulated (Fig 1C). The top 50 genes arranged by p-value are shown in Figure 1D. Among the upregulated genes are several cancer-related transcriptional regulators including PPARD (27) and HMGA1 (28), key regulators of lipid pathways (29), transcription elongation factor SPT6 (SUPT6H), SOAT2, an enzyme involved in lipoprotein and cholesterol regulation, and GPD1, an enzyme that plays a key role in lipid metabolism, are also upregulated in responders. Among the downregulated cancer-related genes we found five involved in mitosis and G2/M DNA replication checkpoint, the kinesin-like proteins KIF11 and KIF15 (30), centromere/centrosome proteins CENPF and CEP55 (31), and the cell cycle regulator protein DLGAP5 (32). Importantly, CEP55 and DLGAP5 are key predictors of antibody response in our graphical model discussed below.

We then performed double contrast analysis of the genes that were significantly differentially changed from baseline to week 2 in the responders vs. non-responders (Fig 1E). The top 50 genes by p-value (Fig 1F) are enriched in immune-related genes. IFNL1 is upregulated in contrast to CD38 and IL12RB2, which are downregulated in responders. Again, selective upregulation of transcriptional and epigenetic regulators in responders is evident; examples include PRDM5, ZNF230, ZCCHC9, ZKSCAN4, and the epigenetic regulator ALKBH3, which demethylates DNA and RNA in cancer cells (33).

### ICOS/ICOSL signaling is differentially associated with response to the MUC1 vaccine

We performed gene set variation analysis (GSVA) of the baseline gene expression data to identify biological pathways regulating the response to vaccination. Vaccine responders displayed significant upregulation of genes involved in the ICOS-ICOSL pathway in T-helper Cells signaling pathway, with increased expression of multiple genes both at baseline and week 2 post-vaccination (Fig 2A and 2B, respectively). At baseline, responders expressed higher levels of ICOSL, IL2RB, and CD4 coreceptor genes, while at week 2 post-vaccination higher levels of the downstream NFKB pathway genes including NFKB2, RELB and RELA were evident.

**Figure 2:**
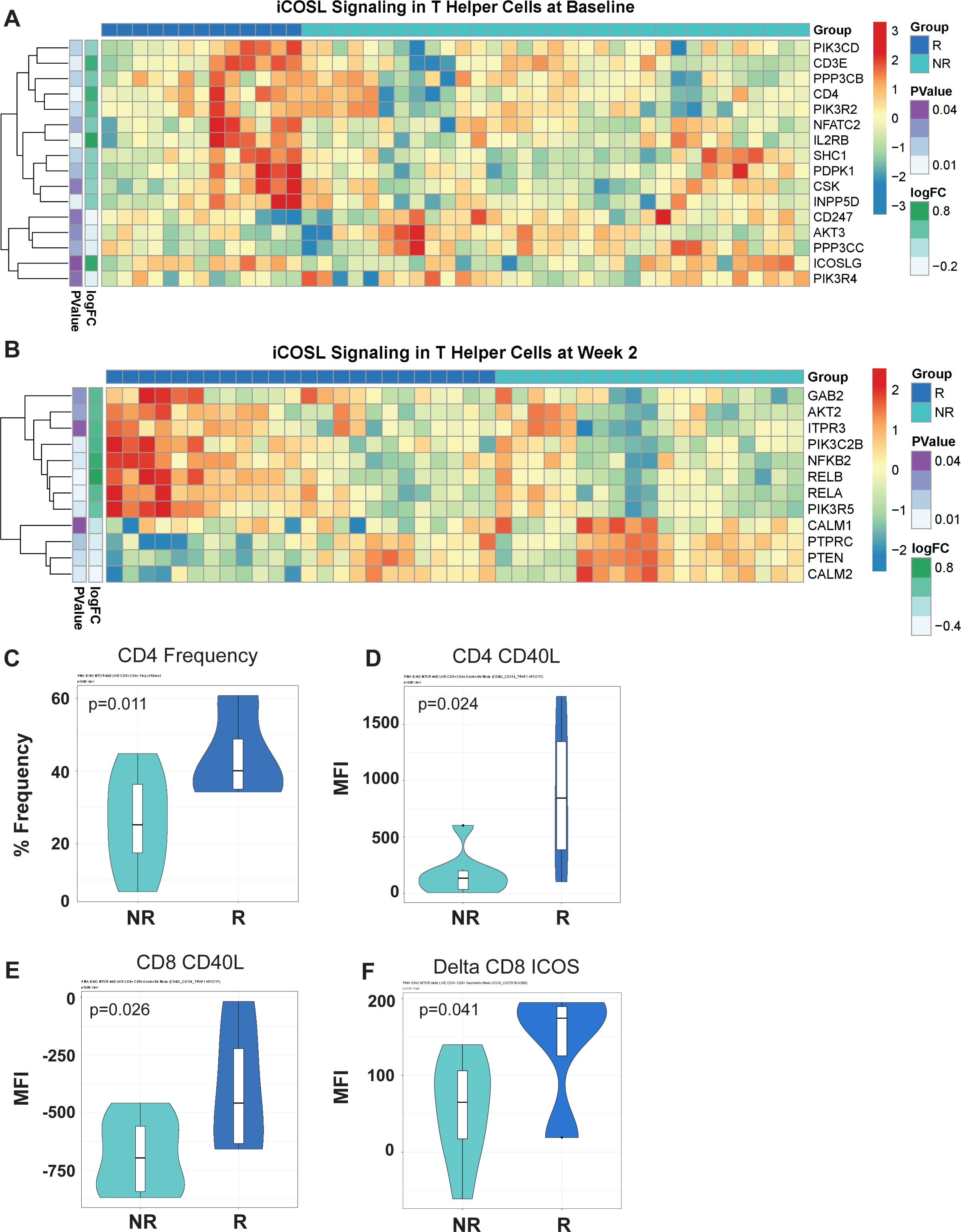
Differentially expressed T-cell fitness signatures in PBMCs from responders and non-responders to MUC1 vaccination is associated with CD4 frequencies and expression of multiple regulators of T-cell help. **(A,B)** iCOSL signaling pathway-related genes are associated with response to MUC1 vaccination at Baseline (A) and Week 2 post-vaccination (B) in MUC1 vaccine responders and non-responders. Heatmaps are organized as in Figure 1. (C) Violin plots of CD4+ T-cell frequencies as determined by flow cytometry. (D-F) Violin plots of CD40L expression on CD4 T-cells (D), CD8 T-cells (E), and the change in ICOS levels on CD8 T-cells (Delta Week 2 vs. Week 0/Baseline) (F) measured by geometric mean fluorescence intensity (MFI).

By flow cytometry, we determined that higher frequencies of CD4 T-cells were present in responders at baseline, potentially related to enhanced ICOSL signaling (Fig 2C). We also detected differences in expression levels of key proteins in this pathway. Higher levels of expression of CD40L were evident in CD4 and CD8 cells (Fig 2D and 2E respectively). Greater increases in ICOS expression were detected post-vaccination inCD8 T-cells of responders. (Fig 2F).

### mTOR signaling is upregulated in responders to MUC1 vaccination

As many components of the ICOS/ICOSL pathway were significantly higher in the responders vs. non-responders, we focused on differences in the mTOR signaling pathway which lies directly downstream of ICOS/ICOSL engagement. Top enriched pathways in high responders vs. non-responder comparisons included PI3K/AKT/MTOR signaling, WNT/beta-catenin signaling and hedgehog signaling (Fig 3A). In contrast, the Myc targets V1 pathway and DNA repair were negatively associated in high responders.

**Figure 3:**
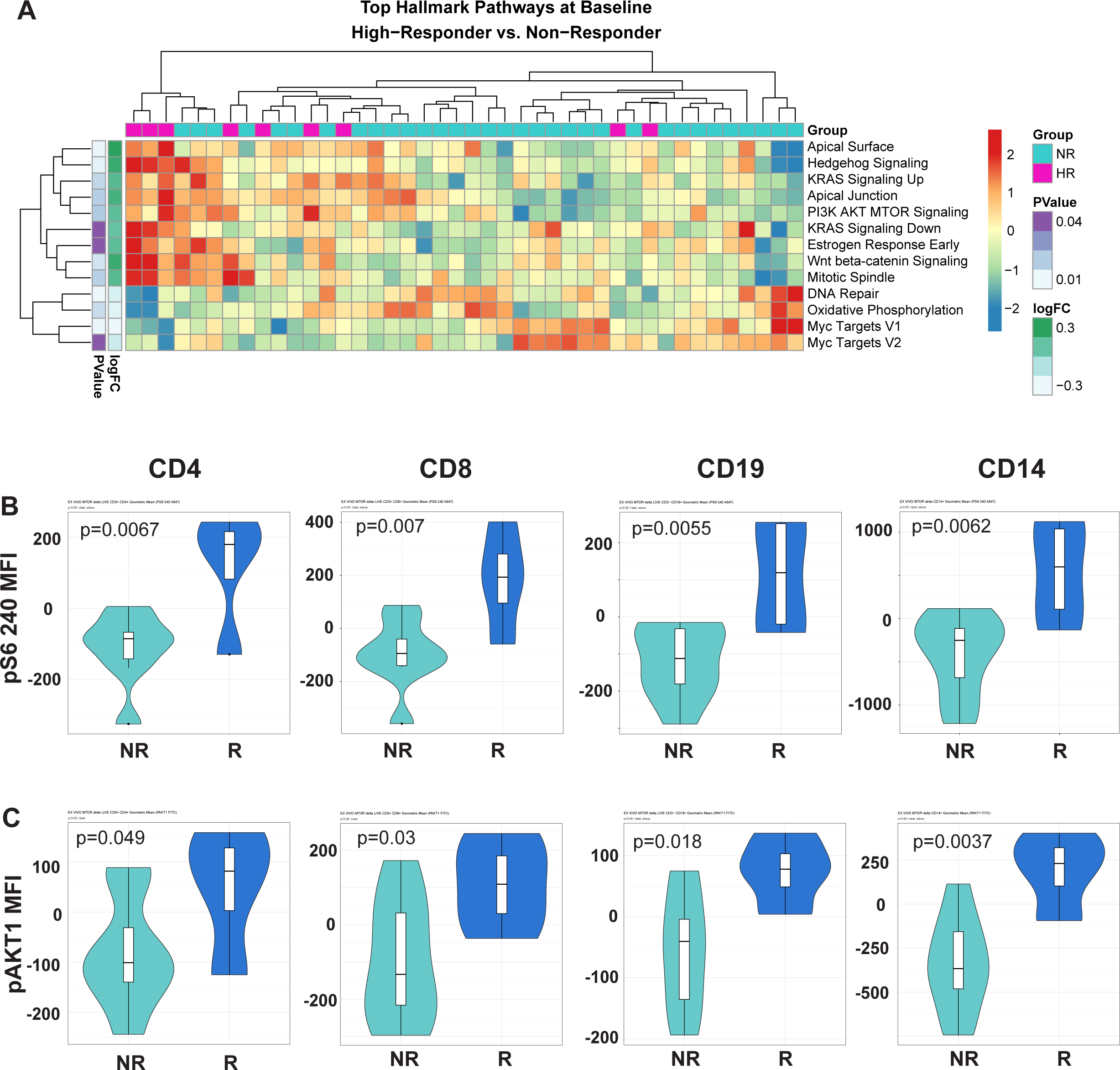
The mTOR signaling pathway is upregulated in enhanced response to MUC1 vaccination. **(A)** Heatmap showing top differentially enriched pathways from the Hallmark Gene Set (MSigDB) in the PBMCs from high responders (HR) vs. non-responders (NR) at Baseline. Group status is indicated in the row above the heatmap as follows: high responders (HR) - pink, while non-responders (NR) - light blue. Z-scored normalized pathway enrichment log 2 fold-change and p values are displayed as in Figure 1. Unsupervised clustering of the samples is indicated at the top of the heatmap, while clustering of the pathways is displayed on the far left. **(B)** Violin plots showing the level of S6 ribosomal protein phosphorylation in the indicated cell subsets (CD4, CD8, CD19/B-cells and CD14/monocytes). **(C)** Violin plots showing the level of AKT1 phosphorylation in the indicated cell subsets (CD4, CD8, CD19/B-cells and CD14/monocytes). For all violin plots, geometric mean fluorescence intensity (MFI) is shown on the y-axis. Responders (R) and non-responders (NR) are designated by dark blue and light blue respectively.

To validate enhanced mTOR signaling in responders, we developed a phosflow-based panel of antibodies (see Materials and Methods) to measure the phosphorylation levels of RPS6, a commonly used readout of mTORC1 activity. There was a greater increase in RPS6 phosphorylation in CD4 and CD8 T-cells, B-cells (CD19), and monocytes (CD14) of responders, an observation validating our finding at the phosphoprotein level (Fig 3B). We also performed intracellular staining targeting the phosphorylated AKT1 kinase upstream of the MTORC1 signaling complex. Similarly, we found a greater increase in AKT1 phosphorylation in responders compared to non-responders.

### B-cell signaling and enhanced antigen presentation signatures are positively associated with response to MUC1 vaccination

As ICOS/ICOSL-mediated signaling promotes fitness of the T lymphocyte compartment, we hypothesized that signaling from the T-cells to the B-cell and APC compartment was also differentially induced. Pathway enrichment analysis of the transcriptomic data revealed significant enrichment of B-Cell Receptor Signaling and PI3K Signaling in B Lymphocytes pathways at baseline (Fig 4A and 4B) and NFκB and CD40 Signaling pathways at baseline and at week 2 post-vaccination (Fig 4C and 4D). CD40 receptor engagement on the surface of antigen presenting cells, such as B-cells, leads to activation of NFκB signaling and enhanced cellular survival and function. Notably, multiple signaling component genes (MAP kinases and Jak3) are significantly upregulated in responders at baseline, followed by upregulation of additional signaling molecules at week two (TRAF1, TRAF3, NFKB1, NFKB2, RELA and RELB).

**Figure 4:**
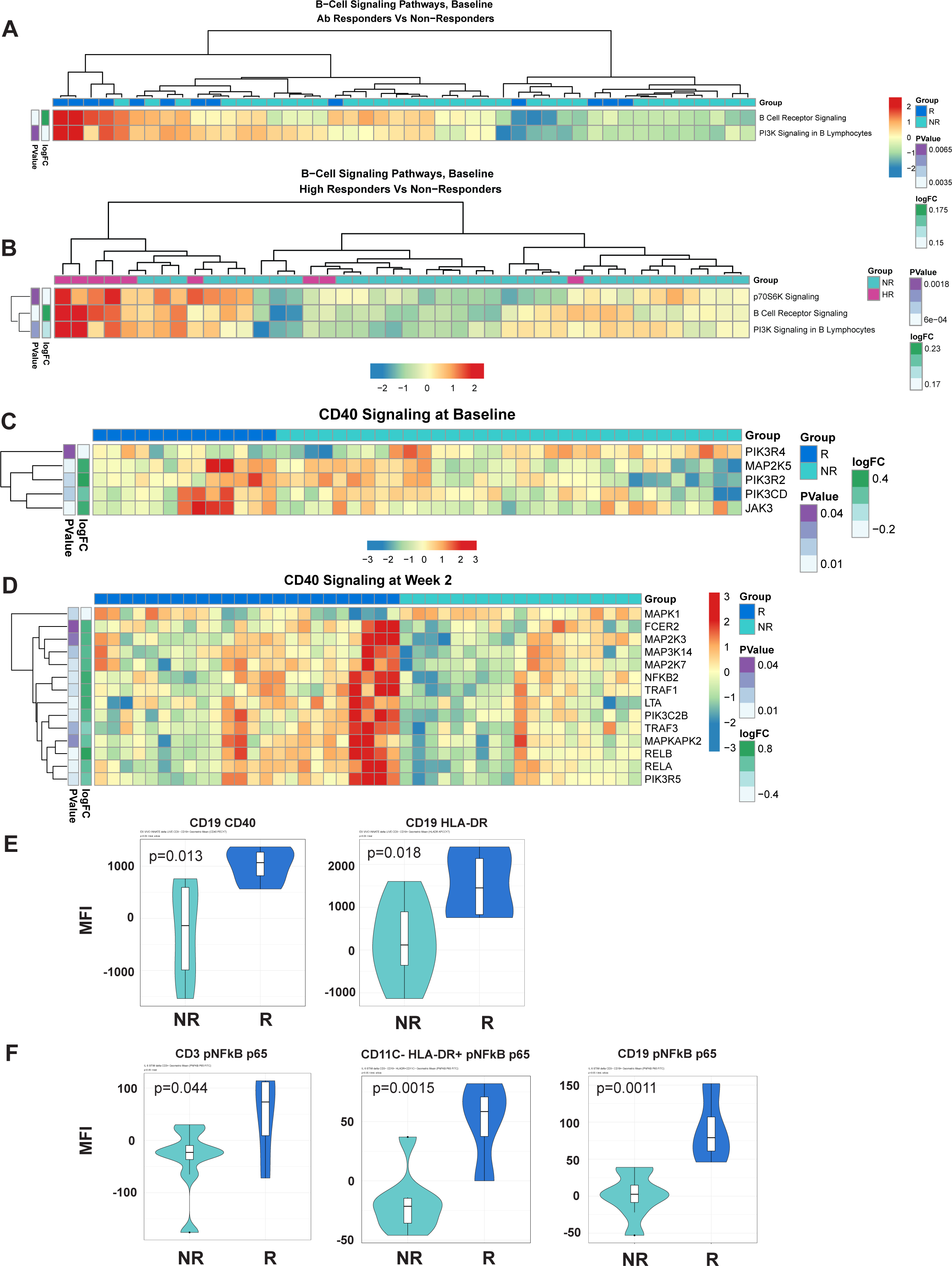
B-cell signaling and NFκB signaling signatures are associated with response to MUC1 vaccination. **(A,B)** Heatmaps showing enrichment of B-Cell Receptor Signaling pathways in the PBMCs from Responders (R) vs. non-responders (NR) at Baseline (A) and high responders (HR) vs. non-responders (NR) at Baseline (B). **(C,D)** Differential gene expression from the CD40 Signaling pathway from PBMCs from R vs. NR at Baseline (C) and Week 2 (D). Heat maps are organized as in Figure 1. **(E)** Violin plots showing expression levels of CD40 and HLA-DR on B-cells (CD19+). **(F)** Violin plots of NFκB complex p65 subunit phosphorylation in T-cells (CD3+), non-DC/non-B antigen presenting cells (CD11C-HLADR+), and B-cells (CD19+). For violin plots, geometric mean fluorescence intensity (MFI) is shown on the y-axis.

We validated increased expression of CD40 and HLA-DR on B-cells (CD19+) in responders (Fig 4E) using flow cytometric analyses. We used an intracellular phosflow panel to detect phosphorylation of the p65 subunit of NFκB. We found increased IL6-induced NFκB signaling via phosphoryation of p65 in T-cells, HLA-DR+ non-B/non-DC APCs and B-cells of responders (Fig 4F), and B-cells of high responders expressing significantly higher levels of HLADR compared to non-responders (Fig 4E).

Finally, we performed gene set variation analysis using the Nakaya et al. vaccine immunogenicity pathway database (34) and determined that a plasmacytoid dendritic cell (DC) signature was already enriched in the PBMCs of responders at baseline (Fig 5A) and further enriched at week 2 post-vaccination (Fig 5B). Vaccine responders showed increased HLA-DR levels on DCs (CD3-, CD19-, HLA-DR+, CD11c+) at baseline (Fig 5C). Responders also showed a greater relative change in CD86 and CD40 expression from baseline to two weeks (Fig 5D-E). Altogether, these results indicate that additional signatures of enhanced antigen presentation are associated with enhance response to MUC1 vaccination.

**Figure 5:**
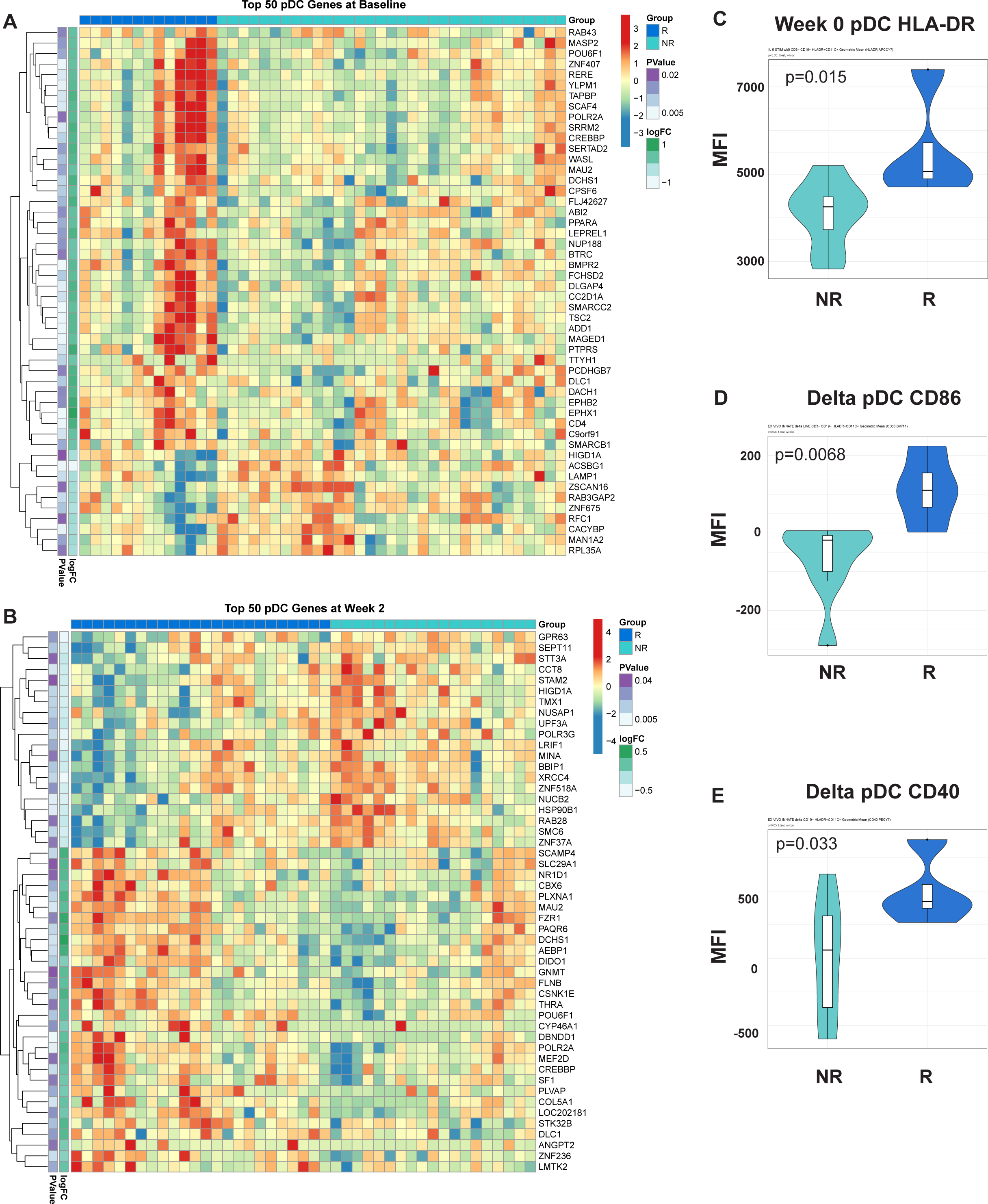
Signatures of enhanced antigen presentation are evident in participants with an enhanced response to MUC1 vaccination. Heatmaps showing enrichment of dendritic cell (DC) specific genes in the PBMCs from Responders (R) vs. non-responders (NR) at Baseline (A) and Week 2 post-vaccination (B). Heatmaps are organized as in Figure 1. Violin plots of HLA-DR expression on DCs at Baseline (Week 0) (C), as well relative change in expression of CD86 (D) and CD40 (E) in these cells. For all violin plots, geometric mean fluorescence intensity (MFI) is shown on the y-axis.

### Six differentially expressed transcripts 2 weeks post-vaccination predict week 12 IgG response to the MUC1 vaccine

Based on evidence of key differences in cell populations and molecular pathways between responders and non-responders at baseline and post-vaccination, we hypothesized that some differentially expressed genes may be useful for patient selection and outcome prediction. We tested this hypothesis by applying LASSO regression and MGM-FCI-MAX (17), a graphical modeling algorithm, to 7,968 transcripts meeting a minimal variance threshold. We first performed a cross-validation experiment (see Supplemental Methods) to assess the ability to predict antibody response to the vaccine at week 12, using the transcriptomic signatures at week 2 post-vaccination. Our model achieved an area under the receiver operating characteristic curve (AUROC) value of 0.741 to predict response vs. non-response (Fig 6A). At a predicted probability threshold of 0.5, the model achieved a sensitivity of 91.7% (22 predicted responders / 24 true responders) and a specificity of 36.8% (7 predicted non-responders / 19 true non-responders). Predicted response odds were correlated with the magnitude of antibody titer at week 12 (R^2^ = 0.209, p<0.001) (Fig 6B), and with the ratio of IgG titer at week 12 versus baseline (R^2^ = 0.147, p=0.015).

**Figure 6:**
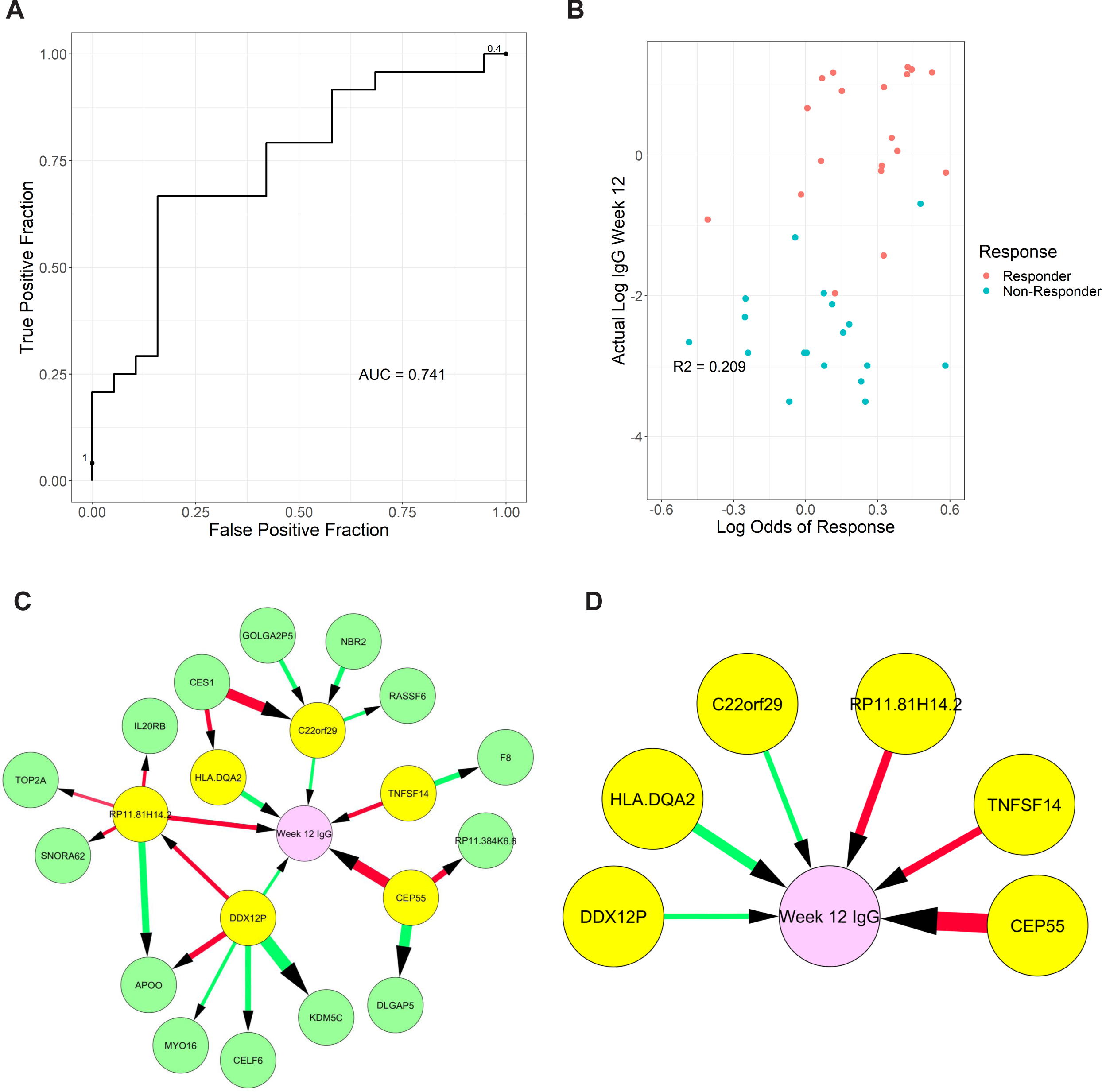
Graphical models of response from transcriptomic data measured two-weeks post-vaccination. (A) Receiver Operating Characteristic curve of response (≥2-fold increase in IgG) using week 2 transcriptome signature. (B) Correlation of predicted response odds with the magnitude of antibody titer at week 12 (C) Full model showing all neighbors and second neighbors of week 12 antibody titer levels (Week 12 IgG), (D) reduced model showing only direct causes of week 12 antibody titer. Color of edge denotes a positive vs negative correlation, and size denotes edge stability.

Next, we used graphical models to determine the variables directly linked to week 12 antibody titer and distinguish them from simple correlates. We produced a full model with all genes selected in the previous cross-validation experiments, which were learned using the entire week 2 dataset (Fig 6C). Finally, we identified 6 genes that are directly linked to week 12 antibody titer: RP11.81H14.2, CEP55, and TNFSF14 (negatively associated) and C22orf29, DDX12P, and HLA-DQA2 (positively associated) (Fig 6D). The role of these transcripts and how they may contribute to the induction of immune response and vaccine efficacy (discussed below) warrants further investigation.

## Discussion

Therapeutic cancer vaccines, tested in numerous clinical trials over several decades, failed to realize the promise generated by the discovery of tumor antigens capable of eliciting humoral and cellular immunity. In most cases, vaccines administered after primary tumor removal failed to boost anti-tumor immunity and prevent tumor recurrence. Ultimately, a greater understanding of the many immunosuppressive forces in the tumor microenvironment helped to explain the reduced efficacy of therapeutic vaccines. These discoveries support preventative vaccines as an alternative approach to cancer vaccination to reduce cancer risk and incidence as they could be applied in the absence of cancer and cancer-induced immunosuppression. The two clinical trials from which we derived the PBMCs, applied this preventative approach by vaccinating individuals without cancer but at high risk for colon cancer due to advanced colonic adenoma diagnosis (13, 14). We expected a vaccine response from most individuals, measured by the production of anti-MUC1 IgG antibodies. It came as a surprise, therefore, that the majority did not respond. However, those who responded had high levels of anti-MUC1 antibodies and established a long-lasting memory response, showing that the vaccine was capable of inducing immunity and response was determined by the individuals receiving the vaccine. Mechanisms underlying this variable response to an apparently efficacious vaccine were not clear.

To address this major knowledge gap with an unbiassed approach, we performed RNA-seq on total PBMCs from participants in the two MUC1 peptide vaccine trials to identify genes and pathways differentially regulated at baseline as well as post-vaccination in the participants that responded versus those that failed to respond to the vaccine. The analysis revealed that vaccine responders at baseline exhibited an enrichment of key pathways governing survival and proliferation in immune cells, such as mTOR and NFKB signaling, as well as increased frequencies of CD4 and CD8 T-cells. There were more memory CD8 T-cells at baseline and week 2 (post MUC1 vaccine) in responders (Fig 2A and 2E). Responders also had more CD4 T-cells at baseline and higher frequencies of memory CD4 T-cells at week 2 (Fig 2A, 2C, 2D). Other immune compartments appeared to differ in favor of responders with higher levels of BCL2 expression at baseline in CD14+ and greater increases in BCL2 expression at week 2 post-vaccination (data not shown). The control of DC longevity by the regulation of BCL2 directly impacts immune responses. Higher levels of BCL2 suggest enhanced survival in the myeloid compartment and consequently better antigen presentation.

The differentially expressed genes and pathways that we have identified in vaccine responders and non-responders pre-vaccination and two weeks post-vaccination are top candidates for early biomarkers of vaccine immunogenicity at week 12. Among the six genes directly linked to MUC1 antibody production at week 12, some have already been identified as diagnostic biomarkers (CEP55, TNFSF14) while the others merit deeper investigation as they hold the potential to enhance our understanding of vaccine response. Overexpression of CEP55 has been observed in numerous cancer cell types, including premalignant lesions of the colon (35), and is a known correlate of poor prognosis (36). Notably, a CEP55 peptide vaccine was proposed for breast and colorectal carcinoma immunotherapy as CEP55 is involved in the PI3K/Akt signaling pathway. TNFSF14, also known as LIGHT, functions as a co-stimulatory factor for the activation of lymphoid cells and modulates T-cell proliferation (37, 38). HLA-DQA2 codes for the alpha chain of the HLA-DQ complex and is primarily involved in antigen presentation (37). Interestingly, HLA-DQ phenotypes have been linked with non-responsiveness to hepatitis B vaccination (39). DDX12P is an m^6^A-associated prognostic pseudogene, correlated with favorable outcomes in patients with head and neck squamous cell carcinoma (40). Furthermore, expression patterns of DDX12P were correlated with anti-tumor response and may regulate immune-involved genes through miRNA targeting. RP11-81H14.2 (LINC02384) is a long intergenic non-coding RNA primarily expressed in TH1 cells (41). Little is known about the function of LINC02384; however, it has been proposed to act as a competitive endogenous RNA of IL2RA and IL7R by reducing available shared regulatory miRNAs (42). C22orf29 (also known as RTL10) may have the capacity to induce apoptosis in a BH3 domain-dependent manner, presumably by engaging the Bcl2 family regulatory network to modulate the intrinsic apoptotic signaling pathway (43). The identification of known diagnostic biomarkers and immunotherapy targets within our predictive genes lends credence to the graphical models utilized in this study.

Given the cancer immunoprevention potential of the MUC1 peptide vaccine response, characterized by a reduction of adenoma recurrence (14), the differentially expressed genes and regulated pathways we identified hold promise as therapeutic targets for vaccine non-responders. While these observations were made on responders and non-responders to the MUC1 vaccine, it is likely that a number of these differentially enriched genes and pathways play a role in other vaccine responses. Many vaccines do not elicit a response in all recipients, such as the yearly flu vaccine which varies in effectiveness between 40% and 60% (44). The selected adjuvant for the MUC1 vaccine, polyICLC, excels at activating dendritic cells to promote type I (innate) immunity (45). Alternative adjuvants may need to be considered for non-responders to the MUC1 peptide adjuvanted with polylCLC. While efforts are often made to improve the vaccine, it also may be important to consider an individual’s incoming immune history to respond to the vaccine. Indeed, numerous research studies have demonstrated a correlation between the immune status prior to vaccination and the subsequent antibody response (46, 47). Overall, individuals that responded to the MUC1 vaccine showed a greater readiness in all the immune compartments to present and respond to antigen. The ability to profile individuals as potential responders or non-responders can aid in the selection of those who benefit most from a particular vaccine. At the same time, understanding the barriers to response in non-responders can inform the development of better vaccine designs suitable for specific immune genotypes and phenotypes.

## Data Availability

The datasets for this study will be found in the Gene Expression Omnibus (GEO) public database with the accession number pending.

## Acknowledgements

We appreciate the tremendous efforts of the team members that conducted the two clinical trials and provided samples for this study, particularly Lisa Boardman, Marcia Cruz-Correa, Ajay Bansal, David Kastenberg, Chin Hur, Lynda Dzubinski, Sharon Kaufman, Luz M Rodriguez, Ellen Richmond, Asad Umar, Eva Szabo, Andres Salazar, John McKolanis, Pamela Beatty, Reetesh Pai, Aatur Singhi, Camille Jacqueline, Riyue Bao, Brenda Diergaarde, Ryan McMurray, Carrie Strand, Nathan Foster, David Zahrieh, and Paul Limburg. We are also indebted to all the trial participants for their commitment to our cancer prevention mission. We thank the Genomics Core at the Lerner Research Institute of Cleveland Clinic and the Genomics and Applied Functional Genomics Cores at Case Western Reserve University for their technical and analytical support.

## Supplemental Material

**Table S1:** Detailed information on antibody panels used for flow cytometric analysis.

Panel 1: Innate Phenotyping
| Company | Cat # | Antibody | Color |
| --- | --- | --- | --- |
| BD | 560114 | STAT3 (pY705) | PerCP-Cy5.5 |
| BD | 558421 | anti-NF-κB p65 (pS529) | A488 |
| BD | 563931 | CD152/CTLA4 | BV786 |
| Biolegend | 305440 | CD86 | BV711 |
| BD | 743008 | CD275/ICOSLG | BV650 |
| BD | 563929 | CD11c | BV605 |
| Invitrogen | L34957 | Live/Dead | Amcyan |
| Biolegend | 318340 | CD56 | BV510 |
| BD | 558122 | CD16 | PB |
| BD | 564303 | CD19 | BUV737 |
| BD | 563546 | CD3 | BUV395 |
| BD | 335796 | HLA-DR | APC Cy7 |
| BD | 557923 | CD14 | A700 |
| BD | 612593 | Anti-ERK1/2 (pT202/pY204) | APC |
| Biolegend | 334321 | CD40 | PE-Cy7 |
| BD | 563601 | BCL-2 | PE-CF594 |
| BD | 555388 | CD11b | PE |

Panel 2: MTOR Signaling
| Company | Cat # | Antibody (Clone) | Color |
| --- | --- | --- | --- |
| BD | 338426 | CD16 | PerCP-Cy5.5 |
| BD | 560048 | anti-Akt1 (PKBa/Akt) | FITC |
| BD | 564058 | CD56 | BV786 |
| BD | 563677 | CD8 | BV711 |
| Biolegend | 304135 | CD45RA | BV650 |
| Invitrogen | Q10008 | CD4 | Q.605 |
| Invitrogen | L34957 | Live/Dead | Amcyan |
| Biolegend | 302242 | CD19 | BV510 |
| BD | 561457 | pS6 (S235/S236) | V450 |
| BD | 564444 | CD14 | BUV737 |
| BD | 565885 | Anti-ICOS (CD278) | BUV395 |
| BD | 563588 | Anti-Human CD154 (CD40L) | APCeFluor780 |
| BD | 557943 | CD3 | A700 |
| BD | 560432 | Anti-S6 pS240 | A647 |
| BD | 562321 | CD19 | PE-CF594 |
| BD | 560684 | Anti-Human CD28 | PE-Cy7 |
| BD | 560285 | p4E-BP1 (T36/46) | PE |

## Supplemental Methods

### Causal Modeling with MGM-FCI-MAX

A Probabilistic Graphical Model (PGM) represents the joint distribution of variables in a dataset as a graph where each node corresponds to a variable and an edge between two nodes, A and B, corresponds to a conditional dependence between A and B given the rest of the variables in the data (48). PGM’s come in two types: directed graphical models rely on additional assumptions to infer cause and effect direction between variables, while undirected graphical models indicate only conditional dependence.

MGM-FCI-MAX is a new method to learn a directed model (17). The algorithm begins by inferring an undirected graphical model using the Mixed Graphical Models (MGM) algorithm (16) and then uses FCI-MAX to determine causal direction. MGM models categorical variables as multinomial and continuous variables as Gaussian with a mean given by a linear regression on all other variables. The full joint distribution of the model is given in Equation 1 below. Here, *x_s_* is the sth of p continuous *y*_*j*_ is the jth of q categorical variables. *β*_*st*_ is the linear interaction term between two continuous variables, and *α_s_* is the continuous node potential. *ρ*_*sj*_ is the edge potential function between continuous and categorical variables, and it takes on one value for each category of the variable *y*_*j*_. Finally, *ϕ*_*rj*_ is the potential function between two categorical variables with a unique value for all combinations of categories of the variables *y_r_* and *y*_*j*_.

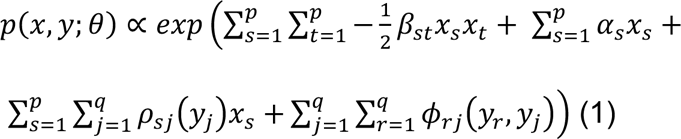

The pseudolikelihood approach was used to optimize the model parameters (49). The pseudolikelihood is the product of the conditional distributions of each variable, and it is a consistent estimator of the goodness of fit of the model to the data. To ensure a sparse graph, edges are penalized via the method proposed in (16) with separate penalty parameters for each edge type: (CC = Continuous-Continuous, CD = Continuous-Discrete, DD= Discrete-Discrete) (Equation 2). Here, *l^~^*(*Θ*) is the negative-log pseudolikelihood and the rest are penalty terms which ensure a sparse model.

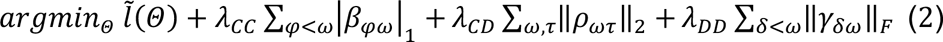

FCI-MAX is used to determine causal directions using the undirected graph as a starting point. FCI-MAX is an extension of the Fast-Causal Inference (FCI) algorithm (50), which is a sound and complete constraint-based algorithm for learning the causal structure of a set of variables in the presence of confounding variables. FCI uses conditional independence tests to rule out unlikely cause and effect relationships. FCI-MAX improves the accuracy of FCI by performing additional tests to more accurately assign orientations, especially in datasets with small sample sizes. The output of the algorithm is a graphical causal model where there are four possible edges. An edge of the form (“A --> B”) suggests that A is a cause of B and B is not a cause of A. An edge (“A <--> B”) suggests that neither A nor B is a cause of the other, that is, a confounding variable causes both. An edge (“A o--> B”) suggests that if there are no latent variables causing both A and B, then A is a cause of B. Finally, an edge of the form (“A o-o B”) suggests that both endpoints are inconclusive. In high dimensional datasets (small sample size, large number of variables) these algorithms are less accurate in inferring causal orientations as they are in inferring the presence or absence of an edge(17).

A likelihood ratio independence test (15) suitable for mixed data was used by FCI-MAX. All three sparsity parameters for MGM (*λ*_*CC*_, *λ*_*CD*_, *λ_DD_*) were set to the default 0.2 and α = 0.1 was used for the independence test threshold for FCI-MAX. MGM-FCI-MAX was run on 100 bootstrap samples of the data, and edges were included in the final model if they appeared in at least 10% of bootstrapped samples.

### Computational Model Development and Evaluation

LASSO logistic regression (23) was used to develop a prediction model (select genes and infer logistic regression coefficients) for a binary outcome of response defined by the clinical trial endpoint (≥2-fold increase in IgG from baseline to week 12), using transcriptomic data measured two-weeks post-vaccination (Week 2 data). To develop and simultaneously evaluate model predictions, a nested leave-one-out cross validation approach was used. Iteratively, each individual sample is used as an evaluation set with the remaining samples used to learn model parameters. On each training set, a LASSO logistic regression was performed with an internal leave-one-out cross validation to choose the optimal sparsity penalty value (λ). The predictions on the single left-out sample in each round of cross validation were then used for downstream analysis.

The Receiver Operator Characteristic (ROC) curve was calculated, and predictive accuracy of the model was measured using the area under the curve (AUC) of response vs. non-response, as well as sensitivity and specificity of predicted probabilities. Feature stability was measured to ensure that models remained similar across different cross-validation iterations.

MGM-FCI-MAX was used to infer the variables directly linked to response, using clinical data (age, sex, and BMI) and those genes selected by LASSO in the week 2 transcriptomic data in at least one of the ten folds. LASSO logistic regression was used to build a predictive model of response in each cross-validation fold. All statistical analysis was performed in R.

